# Automated application of novel gene-disease associations to scale reanalysis of undiagnosed patients

**DOI:** 10.1101/2020.10.16.20212852

**Authors:** Nana E. Mensah, Ataf H. Sabir, Andrew Bond, Wendy Roworth, Melita Irving, Angela C. Davies, Joo Wook Ahn

**Affiliations:** Viapath Genetics Laboratories, Guy’s Hospital, London, UK; Department of Clinical Genetics, Guy’s and St Thomas’ NHS Foundation Trust, London, UK; Birmingham Women’s and Children’s Hospital NHS Trust & Birmingham Health Partners, UK; Faculty of Biology, Medicine and Health, The University of Manchester, Manchester, UK; East Midlands and East of England NHS Genomic Laboratory Hub, Cambridge University Hospitals NHS FT, Addenbrooke’s Hospital, Cambridge, UK

## Abstract

A pressing challenge for genomic medicine services is to increase the diagnostic rate of molecular testing. Reanalysis of genomic data can increase the diagnostic yield of molecular testing for rare diseases by 5.9-47% and novel gene-disease associations are often cited as the catalyst for significant findings. However, clinical services lack adequate resources to conduct routine reanalysis for unresolved cases. To determine whether an automated application could lead to new diagnoses and streamline routine reanalysis, we developed TierUp. TierUp identifies new gene-disease associations with implications for unresolved rare disease cases recruited to the 100,000 Genomes Project. TierUp streams data from the public PanelApp database to enable routine, up-to-date reanalyses. When applied to 948 undiagnosed rare disease cases, TierUp highlighted 410 high and moderate impact variants in under 77 minutes, reducing the burden of variants for review with this reanalysis strategy by 99%. Ongoing variant interpretation has produced five follow-up clinical reports, including a molecular diagnosis of a rare form of spondylometaphyseal dysplasia. We recommend that clinical services leverage bioinformatics expertise to develop automated reanalysis tools. Additionally, we highlight the need for studies focused on the ethical, legal and health economics considerations raised by automated reanalysis tools.

## Introduction

Rare diseases are molecularly defined by clinical DNA sequencing with a diagnostic rate of 29%-49% ^1,2^, hence the majority of patients receive inconclusive results from genetic testing. Disease-causing variants may not be recognised as such due to technical limitations or a lack of sufficient evidence to determine their pathogenicity ^3,4^. Although a patient’s genome sequence is a static resource, our understanding of its relationship to disease is dynamic. Over time, translational research generates new techniques and etiological evidence that may support the discovery of a diagnostic variant ^5^. Reanalysis of inconclusive cases, typically within three years of the initial inconclusive report, has been shown to increase the diagnostic rate by 5.9% to 47% ^5–13^. Successful reanalysis strategies include deeper patient phenotyping ^5,7^, transitioning from singleton to trios ^6,14^, changing assay platforms ^8^, upgrading bioinformatics pipelines and annotation sources ^6,9^, searching for novel gene-disease or variant-disease associations in scientific literature ^5,6,8–10^ and reinterpreting or reclassifying variants ^13^.

Despite having diagnostic utility, reanalysis is not yet routine in clinical laboratories owing to resource constraints and the labour-intensive nature of the practice. For example, upgrading a bioinformatics pipeline can demand 600 hours of work ^7^ and reanalysing just one of approximately 20,000 variants returned per patient from exome sequencing requires several hours of a geneticist’s time ^7,15^. Alternative reanalysis strategies, such as deep phenotyping and transitioning from singleton to trio testing, carry the burden of time spent recontacting patients. In the UK, 16 out of 20 genomic medicine centres report ad-hoc recontacting practices as new tests, new results or new variant classifications become available ^16^, with a similar trend found across Europe ^17^. Clinical laboratories therefore prioritise new referrals rather than incurring the time and labour costs from periodic reanalysis. However, failure to conduct reanalysis leaves patients without diagnoses and may expose clinical services to potential liability ^18^. As professional bodies urge testing laboratories to develop policies for reanalysis ^19^, and clinical genomics moves towards population genome sequencing ^20^, the community will increasingly need to consider automated solutions to reduce the resources required to perform periodic reanalysis ^19^.

A common step in automated reanalysis pipelines is to identify variants that have become associated with genetic disorders following the initial analysis, limiting the number of variants for manual review (variant burden) when combined with additional filtering and prioritisation steps. For example, an automated reanalysis of 240 undiagnosed exomes reduced the variant burden by 93.3% [21]. The authors achieved this by flagging variants with novel gene-disease annotations from either OMIM or PubMed, or novel variant-disease associations from either HGMD or ClinVar. Similarly, the variant burden was reduced by 90% in a partially automated reanalysis of 48 unresolved cases from genome sequencing, driven by a combination of automated re-phenotyping, imputed pathogenicity scores and phenotype-driven ranking ^21^. Efforts to automate reanalysis strategies have increased the diagnostic yield by 4.2%-32% ^21–25^, demonstrating the potential to make significant contributions to patient care. To our knowledge, these pipelines have not been implemented beyond their respective molecular testing centres. One barrier to adoption is the scarcity of simple, open-source implementations that can be shared across laboratories. While the aforementioned studies provide a high-level overview of their methods, substantial bioinformatics expertise is required to reproduce their applications and maintain key data sources over time.

To support the evidence base and guidelines surrounding automated reanalysis, we report the development of TierUp, a lightweight open-source bioinformatics tool, and its contribution to the reanalysis of unresolved rare disease cases. Leveraging clinical bioinformatics expertise within the UK National Health Service, TierUp streamlines the search for new gene-disease associations and re-prioritises variants for review. We demonstrate the utility of two design decisions as yet unseen in automated reanalysis tools: the use of application programming interfaces (APIs) to stream new gene-disease associations in real-time from the curated PanelApp database ^26^, and the use of the Python packaging standard to simplify installation and dependency management. Importantly, we show that our strategy yields additional diagnoses for a rare disease patient cohort with a prior diagnostic yield of 22% ^27^. These findings support the development of community-driven guidelines and policies towards the practice of routine reanalysis in clinical genomics.

## Methods

### Cohort Selection

We selected 948 rare disease patients for reanalysis during November 2019. Eligible patients were participants in the UK 100,000 Genomes Project, recruited through our clinical service at Guy’s and St Thomas’ NHS Foundation Trust (GSTT). Additionally, each case was labelled “unresolved” with an inconclusive diagnostic report.

### Reanalysis strategy

We implemented a two-step reanalysis strategy:

1. Identify variants in genes associated with the patient’s disease subsequent to the initial analysis. These variants were not previously considered candidates as they lacked strong disease associations.
2. Perform standard clinical interpretation of identified variants by applying the ACMG/AMP criteria for classifying variants on a scale from benign to pathogenic^28^.

### TierUp Development

We followed a software development life cycle which included requirements gathering, architecture design, software construction, user acceptance testing and weekly reviews. The result of these activities is TierUp, a command-line tool for identifying variants in genes newly associated with a patient’s disease. TierUp is distributed as a Python package under the JellyPy namespace for tools that interface with the UK national bioinformatics infrastructure provided by Genomics England. Additional documentation relating to the software can be found at https://github.com/NHS-NGS/JellyPy.

TierUp accepts two inputs, a 100,000 Genomes Project case identifier and a configuration file containing API access credentials. Cases recruited to the 100,000 Genomes Project undergo genome sequencing, following which variant filtering is applied to remove failed and common variants, variants with no predicted functional coding impact, and variants that did not segregate with disease within families ^29^. Variants in known disease genes are prioritised for review as determined by gene panels selected at referral from PanelApp, a continuously curated database of crowdsourced expert gene reviews informed by published literature and clinical databases ^26^. TierUp retrieves the latest disease gene lists from the PanelApp API and re-prioritises variants in light of new gene-disease associations, producing a plain text file listing each result.

### Reanalysis

Reanalysis was performed in June 2020 using TierUp version 0.3.0. We installed TierUp on a quad-core laptop with a network download speed of approximately 4MB/s. Case data were passed to TierUp in Javascript Object Notation format and this analysis was distributed across CPU cores using GNU parallel. The linux time command calculated the time taken for TierUp reanalysis.

We combined TierUp outputs from all 948 patients into a single file from which descriptive statistics were calculated using the Python pandas library ^30^ (**Supplementary Materials**).

As a proof of principle, five variants returned by TierUp were interpreted by clinical scientists following standard protocols. These variants were reported to each patient’s referring clinician for further action.

## Results

### Case Demographics

The cohort of 948 rare disease cases undiagnosed by genome sequencing were composed of 368 singletons, 391 trios and 189 with mixed family members. The majority of cases (633/948; 67%) were first analysed in September 2018 and were unresolved for an average of 18 months prior to reanalysis (range 10-20 months).

Cardiovascular disorders (n=232) and neurodevelopmental disorders (n=227) were the most common rare disease referrals. Cases were most commonly referred with a panel of genes associated with intellectual disability (n=254, PanelApp #285).

Collectively, patients carried 564,441 variants for reanalysis with a median of 387 variants per patient (interquartile range = 119 - 739).

### Reanalysis

TierUp processed 564,441 variants in 76 minutes and 44 seconds (Figure 1), of which TierUp flagged 410 variants in genes with newly established disease associations in PanelApp. This equates to 99.93% fewer variants for manual review using this reanalysis strategy. TierUp returned these variants from 121 cases (12.76%) with a median of 1 variant per case (interquartile range = 1 - 2).

**Figure 1.**
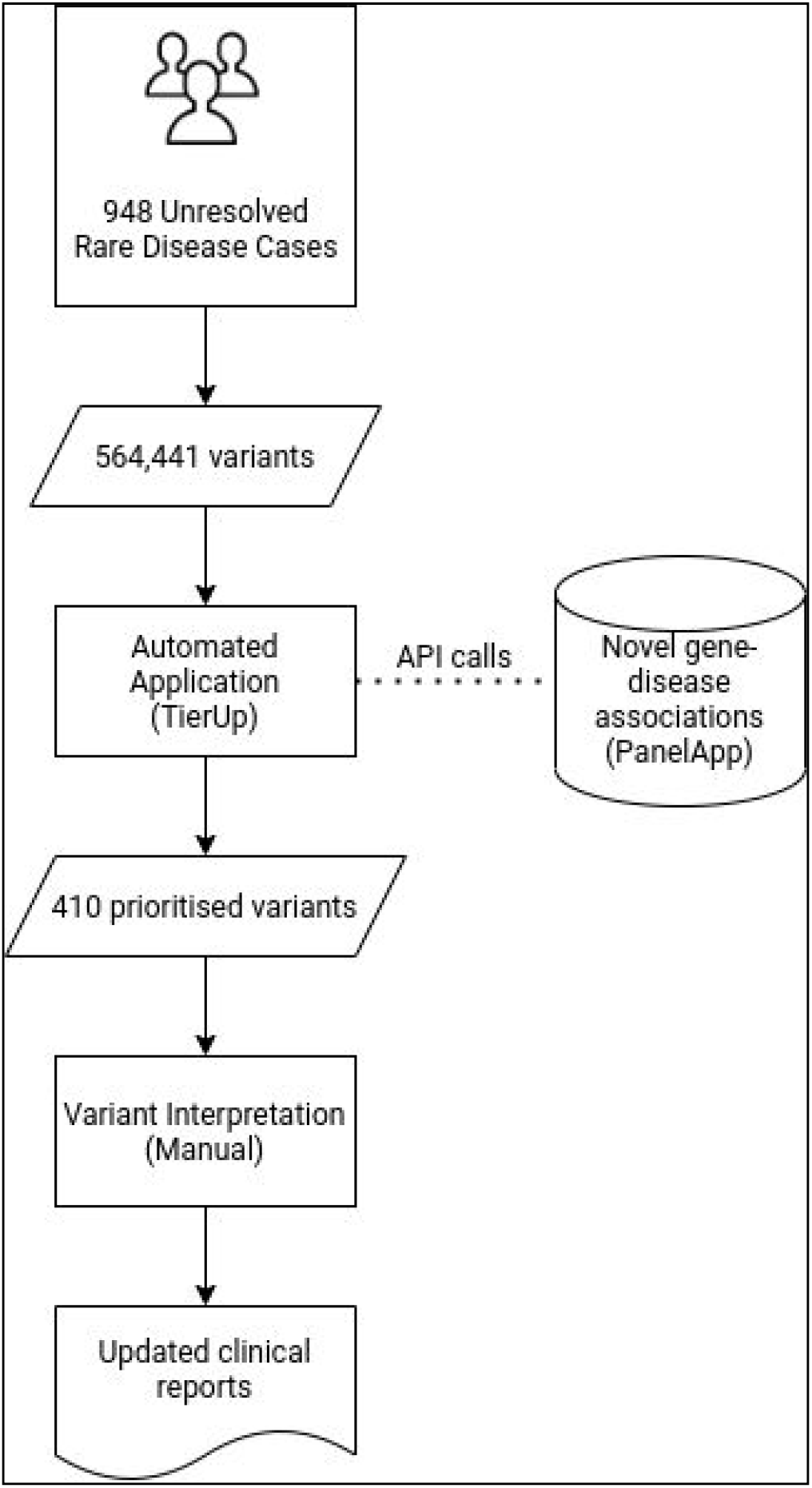
Workflow for TierUp reanalysis of 948 unresolved rare disease cases

Variants were most frequently returned for patients tested with the panels Intellectual Disability (n=73), Congenital Anomaly of the Kidneys and Urinary Tract (n=13), Genetics Epilepsy Syndromes (n=11), Undiagnosed metabolic disorders (n=10), and Skeletal Dysplasia (n=4) (Table 1).

**Table 1.** Patient cohorts with the highest proportion of TierUp results

| Patient Cohort | Percentage of cohort with TierUp results |
| --- | --- |
| Intellectual disability | 28.7 (73/254) |
| Congenital Anomaly of the Kidneys and Urinary Tract (CAKUT) | 27.1 (13/48) |
| Arthrogryposis | 26.7 (4/15) |
| Generalised pustular psoriasis | 25.0 (1/4) |
| Genetic epilepsy syndromes | 20.8 (11/53) |
| Hypogonadotropic hypogonadism | 16.7 (1/6) |
| Limb girdle muscular dystrophy | 15.4 (2/13) |
| Hypertrophic cardiomyopathy - teen and adult | 14.3 (2/14) |
| Clefting | 12.5 (1/8) |
| Anophthalmia or microphthalmia | 12.5 (1/8) |
| Arrhythmogenic cardiomyopathy | 11.1 (1/9) |
| Thoracic aortic aneurysm or dissection | 9.5 (4/42) |
| Undiagnosed metabolic disorders | 9.4 (10/106) |
| Skeletal dysplasia | 8.0 (4/50) |
| Cystic kidney disease | 5.9 (2/34) |
| Retinal disorders | 4.8 (1/21) |
| Skeletal Muscle Channelopathies | 3.7 (1/27) |
| Brain channelopathy | 3.7 (1/27) |
| Congenital muscular dystrophy | 3.2 (1/31) |
| Congenital myopathy | 3.0 (1/33) |
| Early onset dystonia | 2.6 (1/39) |
| Hereditary spastic paraplegia | 2.5 (1/40) |
| Hearing loss | 1.6 (1/63) |
| Mitochondrial disorders | 0.9 (1/107) |

### Variant Interpretation

Five variants prioritised by TierUp have been reported following interpretation, classification and discussion within multi-disciplinary teams (Table 2).

**Table 2.**
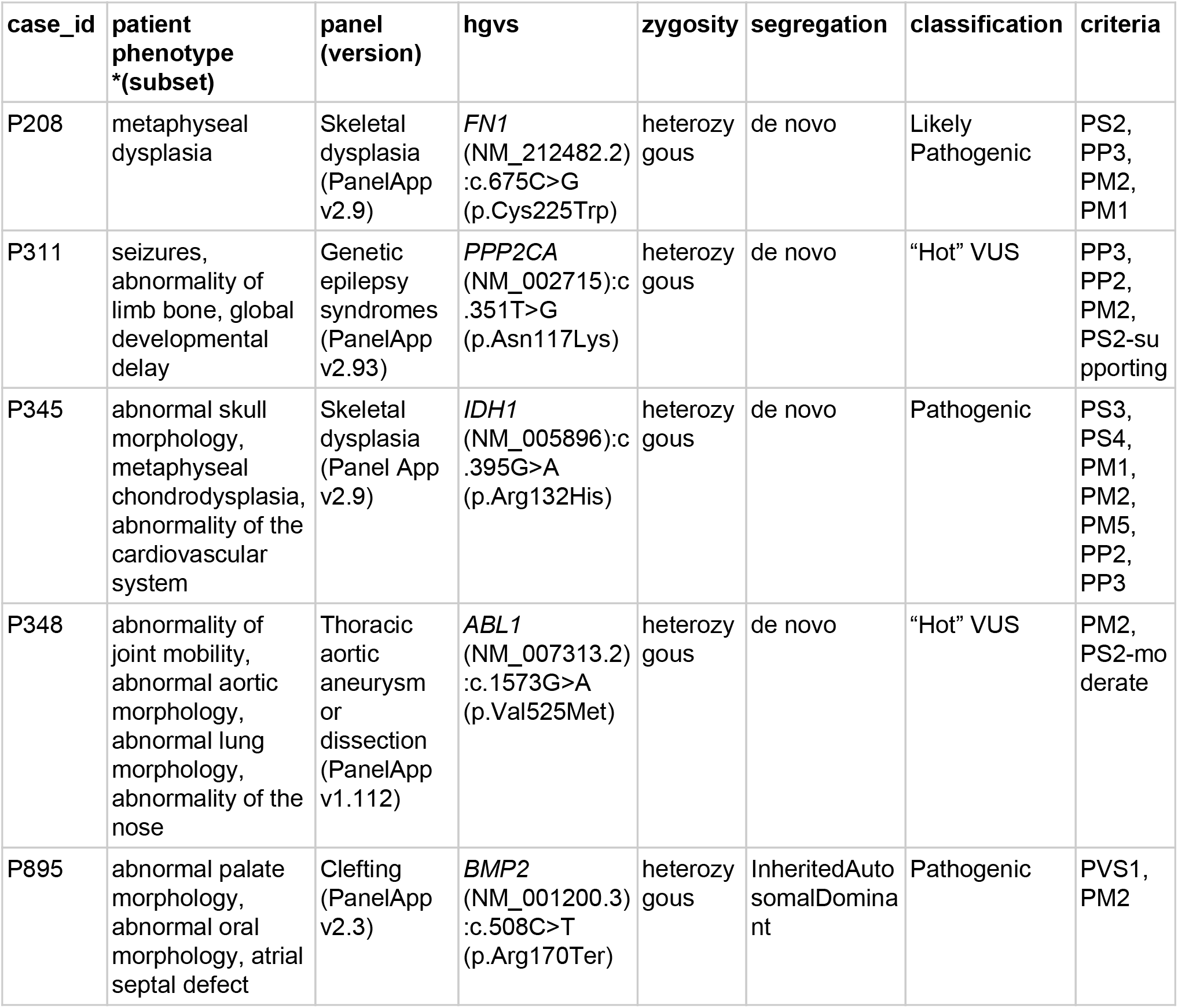
Variants returned by TierUp and reported following variant interpretation

Case P208 obtained a molecular diagnosis as a result of TierUp reanalysis (Figure 2). The patient presented with short stature and facial dysmorphism leading to a preliminary clinical diagnosis of short stature syndrome, Brussels type (OMIM #601350). Genome sequencing was carried out in November 2018 through the 100,000 Genomes Project using the PanelApp skeletal dysplasia panel (PanelApp #309, v1.101), but no significant findings were reported. *FN1*, now a known skeletal dysplasia gene, was added to the panel four months later (PanelApp #309, v1.147). Subsequent reanalysis with TierUp identified the *de novo* heterozygous variant *FN1*(NM_212482.2):c.675C>G (p.Cys225Trp) and the variant was classified as likely pathogenic (PS2, PP3, PM2, PM1). Patient notes and imaging showed phenotypes consistent with pathogenic *FN1* variants, including corner fractures. The patient’s diagnosis has now been reassigned to spondylometaphyseal dysplasia, corner fracture type (OMIM #184255), and the findings were communicated in a clinical case report (submitted for publication).

**Figure 2.**
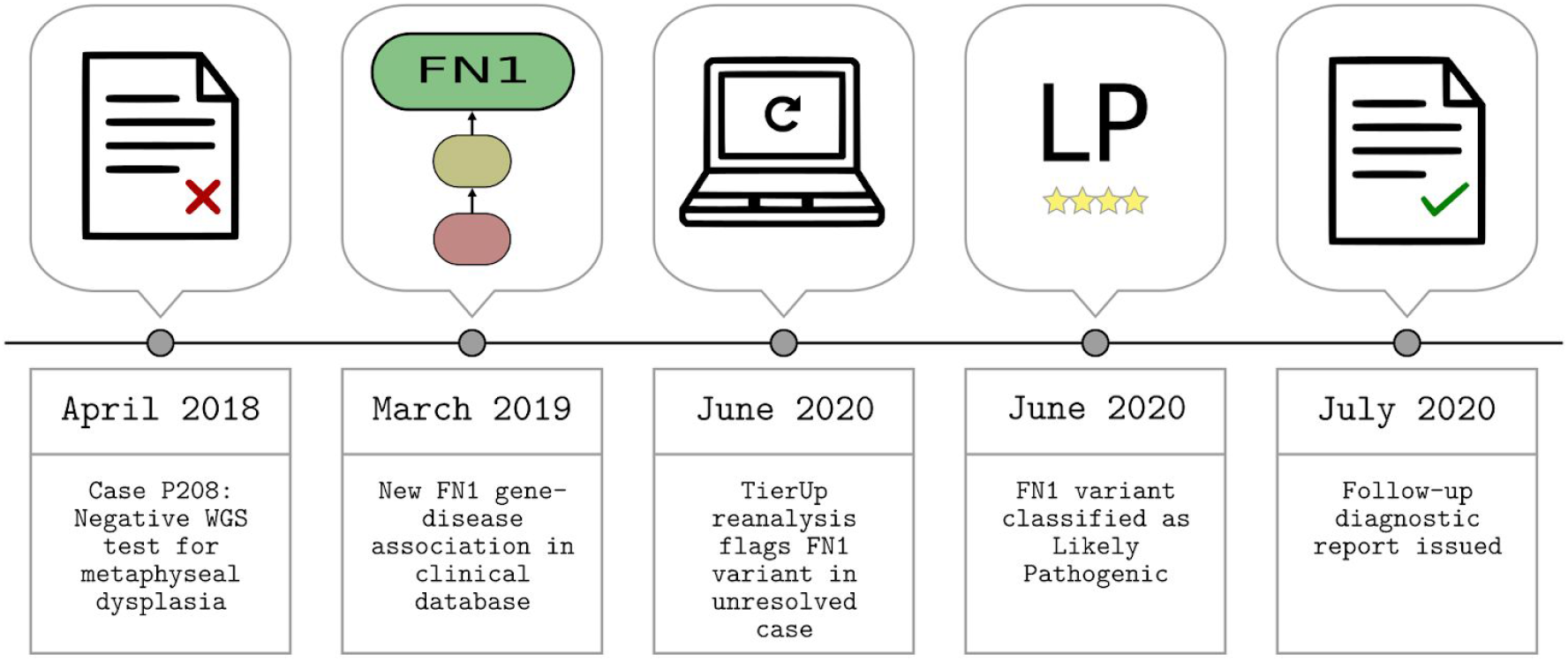
Timeline from negative/inconclusive report to diagnosis for Case P208.

Case P311 obtained an updated clinical report as a result of TierUp reanalysis. The patient presented with seizures and limb abnormalities. Genome sequencing was inconclusive using an early version of the Genetic Epilepsy Syndromes panel (PanelApp Epilepitc Encephalopathy, v2.131). TierUp reanalysis detected the *de novo* variant *PPP2CA* (NM_002715):c.351T>G (p.Asn117Lys) in this patient as *PPP2CA* was later associated with disease in this panel (PanelApp #402, v2.93). *De novo* variants in *PPP2CA* are associated with neurodevelopmental disorders, including intellectual disability, epilepsy and behavioural disorders. We classified the variant as a “hot” variant of uncertain significance (VUS) (PP3, PP2, PM2, PS2-supporting), meaning it has the potential to be upgraded in the future if more evidence supporting pathogenicity becomes available. The variant was later reported at the request of the referring clinician.

Case P345 obtained an updated clinical report as a result of TierUp reanalysis. The patient presented with abnormal skull morphology and metaphyseal chondrodysplasia. Genome sequencing was inconclusive using an early version of the Skeletal Dysplasia panel (Unexplained Skeletal Dysplasias, v1.4). TierUp reanalysis flagged a *de novo* heterozygous variant for review, *IDH1* (NM_005896):c.395G>A (p.Arg132His). This variant has previously been described in unrelated patients with severe chondrodysplasia and is suspected to contribute to this phenotype by reducing the specificity of the NADP+ enzyme encoded by the *IDH1* gene ^31^. The variant was classified as pathogenic (PS3, PS4, PM1, PM2, PM5, PP2, PP3) and a follow-up report was issued.

Case P348 obtained an updated clinical report as a result of TierUp reanalysis. The patient presented with abnormal joint mobility and abnormal aortic, lung and nose morphology. Genome sequencing was inconclusive using the Thoracic Aortic Aneurysm or Dissection panel (PanelApp #1, v1.4). TierUp highlighted the variant *ABL1* (NM_007313.2):c.1573G>A (p.Val525Met) during reanalysis as ABL1 was later promoted for clinical use in this panel (PanelApp #1, v1.8). The variant was classified as a VUS (PM2, PS2-moderate). Although this evidence was insufficient to determine pathogenicity, the disease presentation was highly consistent with *ABL1* disorders and the referring clinician requested that the variant be reported.

Case P895 obtained an updated clinical report as a result of TierUp reanalysis. The patient presented with abnormal palate morphology and atrial septal defects. Genome sequencing using the Clefting Panel (PanelApp #81, v1.34) was inconclusive. The variant *BMP2* (NM_001200.3):c.508C>T (p.Arg170Ter) was prioritised by TierUp as *BMP2* was later associated with disease in this panel (PanelApp #81, v1.39). This heterozygous variant is predicted to introduce a premature stop codon and segregates with affected family members in an autosomal dominant inheritance pattern. *BMP2* haplo-insufficiency has an established association with the craniofacial and cardiac phenotypes observed in case P895 ^32^. The variant was therefore classified as pathogenic (PVS1, PM2) and a follow-up report was issued.

## Discussion

Although reanalysis can yield additional molecular diagnoses for unresolved rare disease cases, existing strategies remain too resource-intensive for clinical services to adopt routinely. Automated reanalysis pipelines can leverage new information in clinical databases to prioritise variants, reduce the burden of variants for review and increase the diagnostic yield from molecular genetic testing ^21,23,24^. We developed TierUp to determine whether an automated application that searches for new-gene disease associations could yield new diagnoses while reducing the resource requirements for reanalysis. We applied TierUp to the reanalysis of 948 rare disease cases which had been unresolved for 18 months on average after undergoing clinical genome sequencing.

Previously, analysts would manually search for new gene-disease associations in various databases such as HGMD, ClinVar and PanelApp, as well as searching the literature. Geneticists would require approximately 1300 hours to manually complete the first step of our reanalysis strategy, assuming that each of the >500,000 variants analysed requires 10 seconds to determine whether it is present in a gene associated with the patient’s disease. In contrast, TierUp processed all cases in less than 77 minutes, suggesting that automated reanalysis is a time-efficient alternative to manual analysis methods.

Whereas systematic manual reanalysis requires us to review over 500,000 variants from our patient cohort, TierUp highlighted 410 candidate variants from 132 cases, hence the burden of variants for manual review was reduced by 99.93%. This supports similar findings from previous studies which achieved a >90% reduction in variant burden using automated pipelines ^21,24^. Collectively, these applications demonstrate that bioinformatics tools can scale reanalysis strategies that rely on updates to existing knowledge bases.

Automated reanalysis pipelines typically require clinical laboratories to maintain local copies of relevant knowledge bases ^21–24^. TierUp is unique in that it leverages the PanelApp API to apply the latest gene-disease associations during reanalysis. With over 700 international reviewers contributing on average 364 reviews per month for curation, PanelApp serves as a dynamic and interoperable source of high quality gene-disease associations ^26^, with each panel update carrying the potential for new diagnoses through automated reanalysis. Applications like TierUp will have increasing relevance to future patients as PanelApp serves as the gene panel repository for genomic medicine services globally ^26,33^.

We present five reported variants from the ongoing interpretation of TierUp results. In particular, case P208 exemplifies the diagnostic potential of this application. This patient was initially diagnosed with short stature syndrome (Brussels type), however no significant variants were reported after clinical exome sequencing and subsequent genome sequencing through the 100,000 Genomes Project. TierUp returned a *de novo* variant in the *FN1* gene which was classified as likely pathogenic. Subsequently, the patient was diagnosed with SMD-FN1 and a case report was submitted for publication (submitted for publication). This molecular diagnosis informs patient management and contributes knowledge to the wider clinical genomics community, both of which may have been delayed indefinitely without TierUp reanalysis.

Ideally, clinicians would rest assured that significant variants in unresolved cases would eventually be brought to their attention by automated pipelines, but unfortunately this ideal is forestalled by the limits of existing tools. For example, TierUp identified diagnostic variants by automating a single reanalysis strategy, however additional diagnoses may be gained through incorporating novel phenotype information and variant-specific annotations, both of which have been automated successfully elsewhere ^21^. Furthermore, our patient cohort underwent genome sequencing, however TierUp reanalysis is limited to single nucleotide variants and indels. Copy number variants and structural variants are of high importance in diagnosing genetic disorders ^34^, hence extending TierUp to these variant types would further improve its diagnostic potential. Finally, although distributing TierUp as a Python package allows other clinical genomic centres to easily perform reanalysis on their own 100,000 genomes patient cohorts, it also demands that these services have adequate bioinformatics support to maintain the software and manage data from consecutive TierUp analyses. A more accessible design would be to offer a reanalysis tool with a web interface, complete with automated variant classification and continuous bi-directional communication of new genomic knowledge and phenotypic updates between the testing laboratory and clinic ^35^.

Automated tools will help realise routine reanalysis but not without measures to address the ongoing ethical, legal and health economic considerations. As the party responsible for data storage and analysis, clinical laboratories may be considered to have a duty to reinterpret variants ^36^. Automated tools may further pronounce this duty however it may still be mitigated by other demanding aspects of reanalysis. For example, variant interpretation, Sanger confirmation and patient recontact remain manual processes, and guidelines stress that laboratories must consider available resources before undertaking reanalysis ^37^.

Furthermore, were routine TierUp reanalysis to become the norm, failure to do so may lead to liability for negligent variant interpretation ^38^. The UK currently follows a common law approach to negligence, however legal duties contingent on reanalysis might emerge as professional guidelines and practice develop ^39^. Finally, the health economics of automated reanalysis must be properly evaluated in order to determine reimbursement and implementation policies. Overcoming these uncertainties should be the focus of future studies exploring the application of automated reanalysis tools.

TierUp demonstrates the potential for automated tools to reduce the variant burden and successfully identify diagnostic variants. TierUp was released in the open-source JellyPy repository in collaboration with clinical bioinformatics services within the NHS. We therefore suggest that clinical services would benefit from the development of further open-source bioinformatics tools for reanalysis, empowering clinical services to address the diagnostic odyssey for rare disease patients.

## Supporting information

Data analysis steps

## Data Availability

All code is available https://github.com/NHS-NGS/JellyPy

## Supplementary Materials

### Materials and Methods

IPython Notebook describing data analysis steps, [available as a PDF](https://drive.google.com/file/d/16cRvnRWafQ0dol0119AQwCJaDNw2IHHE/view?usp=sharing).

### Data availability

Repository containing raw anonymised data and IPython Notebook describing data analysis steps: https://github.com/moka-guys/tierup_v0-3-0_2020

## Acknowledgements

We thank the patients who made this research possible by consenting to the use of data collected as part of their care through the 100,000 Genomes Project and the National Health Service. We thank Aled Jones, Natasha Pinto and Graeme Smith for their technical support during the course of the project. We thank Matthias Janson and Paul Smith for advising on variant analysis. We thank Matthew Welland and Dalia Kasperaviciute from Genomics England for their advice during the software development phase of this study.

The 100,000 Genomes Project is managed by Genomics England Limited (a wholly owned company of the Department of Health and Social Care). The 100,000 Genomes Project is funded by the National Institute for Health Research and NHS England. The Wellcome Trust, Cancer Research UK and the Medical Research Council have also funded research infrastructure.

N.E.M conducted this research while enrolled on the NHS Scientist Training programme which is funded by Health Education England.

## Author Contributions

Conceptualization: J.W.A, N.E.M; Formal Analysis: N.E.M; Software: N.E.M, A.B.; Supervision: J.W.A, A.D.; Writing - original draft: N.E.M; Writing - review and editing: N.E.M, J.W.A, A.H.S., M.I., W.R., A.B., A.H.S..

