## Supplementary material for "Automated application of novel gene-disease associations to scale reanalysis of undiagnosed patients": Data analysis steps

### Supplementary Materials

Reanalysis of unresolved rare disease cases using TierUp version 0.3.0.

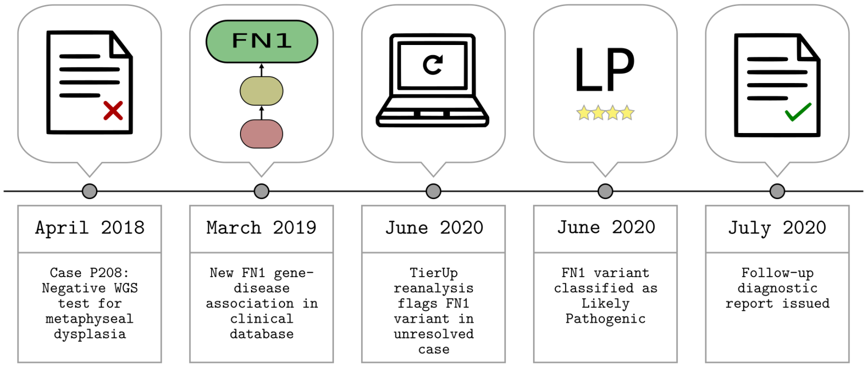


Figure 1: TierUp CaseP208 Timeline

#### Source Data

- db.csv - Merged TierUp results for 948 anonymised cases
- Metadata parsed from TierUp input files using custom scripts:
  - cohort.csv - Number of patients recruited per case
  - rd__group__referrals.csv - Rare disease group referrals from all cases

#### TierUp Reanalysis

TierUp takes GEL interpretation request JSON files as input. These files contain variants prioritised by the GEL tiering pipeline during the initial analysis. TierUp can also download the input files given a case identifier, provided the user is connected to the NHS Health and Social Care Network and has valid GeL CIP-API credentials.

We processed all unresolved cases using the following command: > time parallel tierup -c config.ini -j {} ::: *.jsons &> results.txt

This created `*.tierup.csv` files with the results for each case. Additionally, `results.txt` reported the time taken to process 948 cases on this machine:

```
real    76m43.785s
user    140m17.023s
```

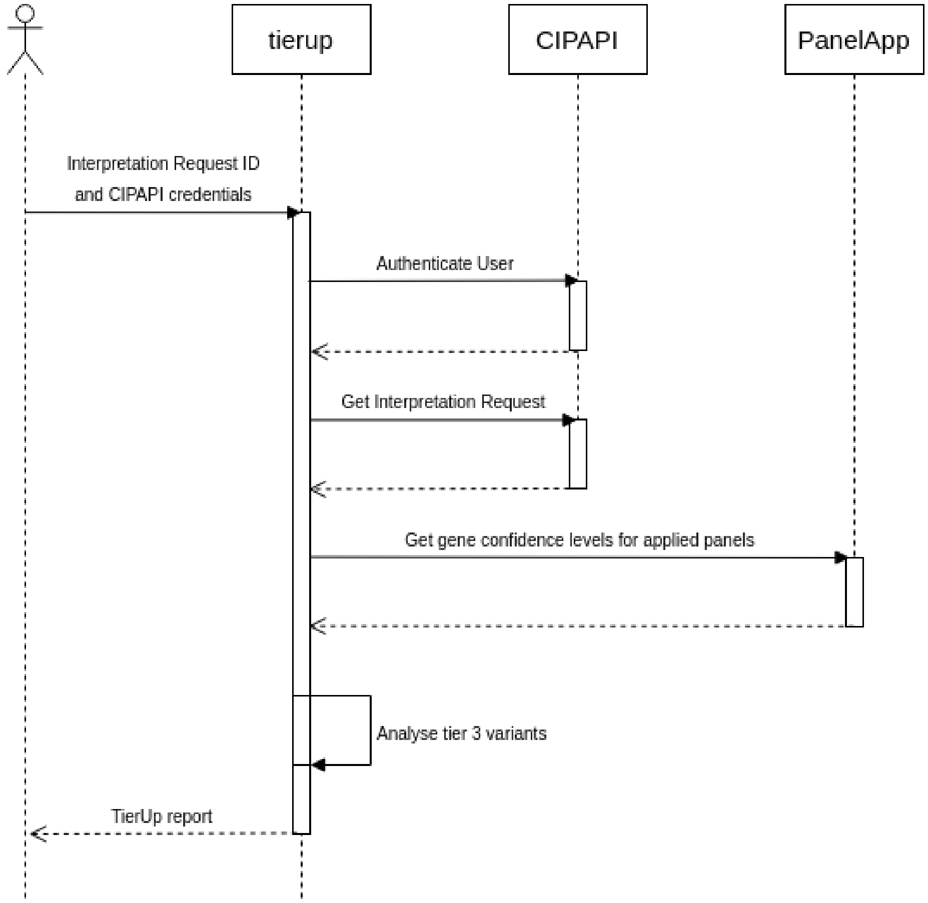


Figure 2: TierUp UML Diagram

```
sys 5m1.144s
```

#### Creating db.csv

We combied TierUp reanalysis results files into a single CSV file using csvstack. Patient identifiers were pseudonymed and any fields that were not required for publication were dropped.

#### Analysis

Descriptive statistics are calculated from db.csv and accompanying files as recorded in the analysis notebook.

#### Files

The `/files` directory contains figures and data that were produced manually.

### tierup_results

September 2, 2020

[3]:
```python
# Imports
import re
import pandas as pd

# Functions
#  result_filter filters db.csv for TierUp signficant hits only. These are high␣
↪and moderate impact variants (tierup tier_1/tier_2)
#  within genes previously not known to be disease causing (gel TIER3)
result_filter = lambda x: x[(x.tier_gel == 'TIER3') & ((x.tier_tierup ==␣
↪'tier_1') | (x.tier_tierup == 'tier_2'))]
```

#### 0.1 Case Demographics

##### 0.1.1 Sample Counts

[4]:
```python
_case_count = pd.read_csv('../data/cohort.csv')

print(str(_case_count.shape[0]) + " samples")

case_count = _case_count.samples.value_counts()
print(case_count)

print(sum(case_count.loc[case_count.index >= 3]))

print(_case_count.trio_bool.value_counts())
```

```
948 samples
1    368
3    364
2    154
4     53
5      7
7      2
Name: samples, dtype: int64
426
false      557
```

```
is_trio    391
Name: trio_bool, dtype: int64
```

##### 0.1.2 Referrals

[5]:
```
referrals = pd.read_csv('../data/rd_group_referrals.csv')
referrals.group.value_counts()[0:6]
```

[5]:
```
Cardiovascular disorders                          232
Neurology and neurodevelopmental disorders        227
None                                              117
Tumour syndromes                                  100
Renal and urinary tract disorders                  87
Dermatological disorders                           48
Name: group, dtype: int64
```

368 singletons, 426 with 3 or more family members, 391 trios

##### 0.1.3 Panels Applied

[6]:
```
df = pd.read_csv('../data/db.csv', usecols=['#id','tu_panel_name']).
 ↪drop_duplicates()
df.tu_panel_name.value_counts()[:10]
```

[6]:
```
Intellectual disability                     254
Familial hypercholesterolaemia              120
Mitochondrial disorders                     107
Undiagnosed metabolic disorders             106
Hearing loss                                 63
Genetic epilepsy syndromes                   53
Skeletal dysplasia                           50
CAKUT                                        48
Familial breast cancer                       44
Rare multisystem ciliopathy disorders        42
Name: tu_panel_name, dtype: int64
```

##### 0.1.4 Time since initial analysis

[7]:
```
# df datetime
_dfdt = pd.read_csv('../data/db.csv', usecols=['#id','created_at',␣
 ↪'tu_run_time'])
dfdt = pd.DataFrame(
    zip(
        _dfdt['#id'],
```

```
        _dfdt['created_at'].apply(pd.Timestamp).apply(lambda x: x.date()),
        _dfdt['tu_run_time'].apply(pd.Timestamp).apply(lambda x: x.date())
    ),
    columns=_dfdt.columns
)

dfdt = dfdt.groupby(list(dfdt.columns)).count().reset_index()
# Add selection date
dfdt['selection_date'] = pd.Timestamp(pd.Timestamp('01 November 2019').date())
# Convert all dates to timestamp to get time differences
dfdt.set_index('#id', inplace=True)
dfdt = dfdt.apply(pd.to_datetime)
dfdt['days_old_at_selection'] = dfdt.selection_date-dfdt.created_at
dfdt['days_old_at_runtime'] = dfdt.tu_run_time-dfdt.created_at

# Confirm that there is one time record per case
assert _dfdt['#id'].unique().shape[0] == dfdt.shape[0]

dfdt.days_old_at_selection = dfdt.days_old_at_selection.apply(abs).apply(lambda␣
↪x: x.days)
dfdt.days_old_at_runtime = dfdt.days_old_at_runtime.apply(abs).apply(lambda x:␣
↪x.days)
months_old_at_runtime = dfdt.days_old_at_runtime.describe()/31
months_old_at_runtime
```

[7]:

```
count    30.580645
mean     18.412856
std       2.655159
min       9.935484
25%      16.830645
50%      20.451613
75%      20.483871
max      20.483871
Name: days_old_at_runtime, dtype: float64
```

##### 0.1.5 Variants to reanalyse

```
[8]: df = pd.read_csv('../data/db.csv', usecols=['#id'])
df['counts'] = 1

variants = df['#id'].shape[0]
print(f'Variants to analyse: {variants}')

case_summary = df.groupby('#id').sum().counts.describe()
print(f"Median variants per case: {case_summary['50%']}\n IQR:␣
↪{case_summary['25%']} - {case_summary['75%']}")
```

```
Variants to analyse: 564441
Median variants per case: 384.0
 IQR: 118.5 - 739.25
```

#### 0.2 Reanalysis

##### 0.2.1 Variants per case

```
[9]: df = pd.read_csv('../data/db.csv', usecols=['#id', 'tier_tierup', 'tier_gel'])

# hmi = High and moderate impact variants returned by TierUp where original
hmi = result_filter(df)

sig_cases = hmi['#id'].nunique()
sig_variants = hmi.shape[0]
print(f'TierUp returned {sig_cases} cases with {sig_variants} significant␣
↪variants')
var_perc = (df.shape[0] - sig_variants) * 100 / df.shape[0]
print(f"This resulted in {round(var_perc, 2)} % fewer variants for review")
```

```
TierUp returned 121 cases with 410 significant variants
This resulted in 99.93 % fewer variants for review
```

```
[10]: hmi = hmi.copy() # Create copy to stop warnings when setting data in slice of␣
↪dataframe
hmi['counts'] = 1
hmi_summary = hmi[['#id','counts']].groupby('#id').sum().describe()

print(f'Cases with significant variants had {hmi_summary.loc["50%"][0]} '
      f'median variants per case (IQR {hmi_summary.loc["25%"][0]}-{hmi_summary.
↪loc["75%"][0]})')
```

```
Cases with significant variants had 1.0 median variants per case (IQR 1.0-2.0)
```

##### 0.2.2 Patient Cohorts

[11]:
```
# What were the top 10 patient cohorts?

df = pd.read_csv('../data/db.csv', usecols=[
    '#id', 'tier_tierup', 'tier_gel', 'tu_panel_name'
    ],low_memory=False)

hmi = result_filter(df)

# ipa = case counts for each intial panel applied
#  Note that panels applied differs slightly as
_ipa = df[["#id","tu_panel_name"]].drop_duplicates()
ipa = _ipa.tu_panel_name.value_counts()

# tpa = case counts for tierup results panels
_tpa = hmi[['#id', 'tu_panel_name']].drop_duplicates()
tpa = _tpa.tu_panel_name.value_counts()


# Combine to create table for patient cohort report
pac = pd.concat([ipa, tpa], axis=1)
pac.columns = ['n', 'tierup_variants']
pac['percs'] = round(pac.tierup_variants * 100 / pac.n, 1)
pac.sort_values('percs', ascending=False, inplace=True)
pac.to_csv('../results/patient_cohort_tierup_variants.csv', index=True)
pac.head(n=10)
```

[11]:

| | n | tierup_variants | percs |
|---|---|---|---|
| Intellectual disability | 254 | 73.0 | 28.7 |
| CAKUT | 48 | 13.0 | 27.1 |
| Arthrogryposis | 15 | 4.0 | 26.7 |
| Generalised pustular psoriasis | 4 | 1.0 | 25.0 |
| Genetic epilepsy syndromes | 53 | 11.0 | 20.8 |
| Hypogonadotropic hypogonadism | 6 | 1.0 | 16.7 |
| Limb girdle muscular dystrophy | 13 | 2.0 | 15.4 |
| Hypertrophic cardiomyopathy - teen and adult | 14 | 2.0 | 14.3 |
| Anophthalmia or microphthalmia | 8 | 1.0 | 12.5 |
| Clefting | 8 | 1.0 | 12.5 |

##### 0.2.3 Varaints Classified and Reported to Date

[20]:
```
hmi = result_filter(pd.read_csv('../data/db.csv'))

case_gene = [
    ('P208', 'FN1'),
```

```python
    ('P311', 'PPP2CA'),
    ('P345', 'IDH1'),
    ('P348', 'ABL1'),
    ('P895', 'BMP2')
]

case_gene_filter = lambda df, case, gene: df[(df['#id'].str.contains(case)) &␣
↪(df['pa_gene'].str.contains(gene))]
case_gene_dfs = [
    case_gene_filter(hmi, case, gene) for case, gene in case_gene
]

variants_reported = pd.concat(case_gene_dfs).drop(142983) # Drop index of␣
↪additional PPP2CA variant
variants_reported['chr_ref_alt'] =␣
↪variants_reported[['chromosome','reference','alternate']].apply(lambda x:␣
↪",".join(x), axis=1)
variants_reported.to_csv('../results/variants_reported.csv', index=False)
variants_reported[['#id', 'chr_ref_alt',␣
↪'pa_gene','zygosity','segregation','penetrance','tu_panel_name','tu_panel_version']]
```

[20]:

```
         #id chr_ref_alt pa_gene      zygosity                 segregation  \
63471   P208       2,G,C     FN1  heterozygous                      deNovo
142984  P311       5,A,C  PPP2CA  heterozygous                      deNovo
171917  P345       2,C,T    IDH1  heterozygous                      deNovo
172791  P348       9,G,A    ABL1  heterozygous                      deNovo
533770  P895      20,C,T    BMP2  heterozygous  InheritedAutosomalDominant

        penetrance                          tu_panel_name  tu_panel_version
63471     complete                     Skeletal dysplasia             2.900
142984    complete            Genetic epilepsy syndromes             2.930
171917    complete                     Skeletal dysplasia             2.900
172791    complete  Thoracic aortic aneurysm or dissection             1.112
533770  incomplete                               Clefting             2.300
```
